# Determinants of quality Antenatal Care utilization in Kenya: insights from the 2022 Kenya Demographic and Health Survey

**DOI:** 10.1101/2024.06.19.24309209

**Authors:** John Baptist Asiimwe, Angella Namulema, Quraish Sserwanja, Joseph Kawuki, Mathius Amperiize, Earnest Amwiine, Lilian Nuwabaine

**Affiliations:** School of Nursing and Midwifery, Aga Khan University, Kampala, Uganda Email: (JBA); (LN); Mbarara Regional Referral Hospital, Mbarara, Uganda; Programs Department, Relief International, Khartoum, Sudan Email: (QS); Program in Public Health, Department of Family, Population, & Preventive Medicine, Stony Brook, University, Stony Brook, NY, USA Email: (JK); Faculty of Medicine, Mbarara University of Science & Technology, Mbarara, Uganda Email: (EA); Infectious Diseases Institute, Kampala, Uganda Email: (MA)

**Author notes:** Corresponding authors John Baptist Asiimwe School of Nursing and Midwifery, Aga Khan University, Kampala, Uganda; Lilian Nuwabaine School of Nursing and Midwifery, Aga Khan University, Kampala, Uganda /700386588.

**Keywords:** Quality, Antenatal Care, Kenya, Women, factors, determinants, association

## Abstract

**Introduction:** One of the most important strategies to lower mother and newborn fatalities worldwide is providing quality Antenatal care (ANC). The utilization of quality ANC by women of reproductive age and associated factors remains unclear in many developing countries. Therefore, the purpose of this study was to determine the factors associated with the utilization of quality ANC in Kenya.

**Methods:** We analyzed Secondary data from the Kenya Demographic Health Survey (KDHS) 2022, which included 11,863 women. Participants were selected using a two-stage stratified sampling design. Using SPSS, version 20, univariate and multivariable logistic regression was used to analyze the data.

**Results:** Of the 11,863 women, 61.2% (95% CI: 59.7-62.6) received quality ANC. Older mothers (aged 20–34) had a 1.82 (95%CI: 1.15-2.87) times higher likelihood of receiving quality ANC when compared with younger mothers (15–19 years old). Participating mothers who had attended 4 or more ANC visits were 1.42 (95%CI: 1.14-1.79) times more likely to receive quality ANC than those who attended 3 or fewer visits. Comparing participants with and without media access, those with media access were 1.47 (95%CI: 1.06-2.03) times more likely to receive quality ANC. Furthermore, the likelihood of receiving quality ANC was 1.93 (95%CI: 1.21-3.08) and 1.44 (95%CI: 1.01-2.06) times higher for participants in the richest and richer quintiles, respectively, than for those in the poorest quintile. On the contrary, the odds of receiving quality ANC were 0.25 (95%CI: 0.15-0.31) to 0.64 (95%CI: 0.44-0.92) times lower for participating mothers from all other Kenyan regions than for those from the coastal region. Participants whose husbands or partners made decisions for them to seek healthcare, compared with those who made decisions independently were 0.74 (95%CI: 0.58-0.95) times less likely to receive quality antenatal care.

**Conclusion:** The study revealed that about 60% of mothers received quality ANC. Several factors associated with quality ANC were identified: age, region, maternal education, health-seeking decision-making, access to media (TV), time to the health facility, ANC visits, and ANC providers (doctor and nurse/midwife/clinical officer). Maternal health improvement programs should prioritize promoting access to education for girls. Additionally, interventions should focus on promoting shared decision-making and autonomy in healthcare-seeking behaviors among pregnant women and their partners, increasing access to care provided by skilled health workers, and addressing regional disparities in healthcare delivery.

## Introduction

Quality Antenatal care (ANC) encompasses assessments and treatments provided by licensed medical practitioners to pregnant adolescent girls and women to maintain the health of the mother and child (Akter et al., 2023; Kare, Gujo, & Yote, 2021). The absence of quality ANC can have severe consequences, which lead to increased risks of maternal mortality and morbidity, as well as adverse outcomes for the newborn, including stillbirth, low birth weight, and preterm birth (World Health Organization, 2019). Quality ANC is paramount in averting maternal mortality by addressing pregnancy-related complications, emphasizing its significance in improving maternal health outcomes worldwide (Modibia, 2020; Charles Wembonyama Mpoy, 2022; Ajibola Idowu, 2022).

In sub-Saharan Africa, where maternal mortality rates are highest, ANC coverage remains low, with only 65% of pregnant women accessing ANC services and information about the quality of ANC remains scanty (WHO, 2019). However, the quality of ANC is suboptimal in some of these countries with Ethiopia having a prevalence of 31.3%, Uganda at 61.4%, and Nigeria at 45% (Modibia 2020, Towongo et al., 2023 & Ajobola Idowu 2022).

Factors influencing the quality of ANC span socio-economic, demographic, obstetric, antenatal, and facility-related domains (Tuncalp et al., 2016; Pell et al., 2016; Say et al., 2014). Numerous influencing factors of ANC quality, include age, socio-economic status, education level, partner support, urban residence, media exposure, facility choice, and ANC visit frequency, among others (Smith et al., 2020; Garcia et al., 2021; Johnson et al., 2019; Lee & Kim, 2023; Wang et al., 2022; Gupta & Sharma, 2023; Nguyen et al., 2020). Moreover, early initiation of ANC visits, adherence to the recommended visits, and the category of the ANC health providers are linked to higher ANC quality (Ijeoma Nkem Okedo-Alex, 2019; Afulani, 2015).

In Kenya, the maternal mortality rate stands at approximately 342 deaths per 100,000 live births, and the newborn mortality rate is about 19 deaths per 1,000 live births, indicating a significant maternal and child health challenge (UNICEF 2023). In Kenya, the Ministry of Health has made efforts to improve ANC coverage and quality through various initiatives, including the implementation of ANC guidelines, training of healthcare providers, and community sensitization programs (Ministry of Health Kenya, 2021). However, the prevalence of quality ANC, as defined by adherence to ANC guidelines and the provision of comprehensive care, remains unclear and may vary across facilities and regions in Kenya (Ministry of Health Kenya, 2021).

Several studies in Kenya examined the subnational disparities in antenatal care utilization and factors influencing the utilization of focused antenatal care services (Wairoto et al., 2020 & Chorongo et al., 2016). These studies, however, did not provide a thorough understanding of how these parameters interact and affect the quality of ANC usage, at a national level. Additionally, given the high maternal and newborn mortality rates, gaps in the quality of ANC services received by women during their pregnancies, may still exist.

Therefore, to shed light on the prevalence and factors that influence the quality of ANC services in Kenya, we used data from the Kenya Demographic Health Survey conducted in 2022. This study may inform interventions to enhance ANC service delivery and promote positive maternal health outcomes in sub-Saharan Africa.

## Methods

### Sampling design, Data collection, and Source

Data from the 2022 Kenya Demographic and Health Survey (KDHS), which used a two-stage stratified sampling design, was used in this study. The first stage involved selecting 1692 enumeration areas (EAs) or clusters from a master sample frame of 129,067 EAs from the 2019 Kenya population and housing census using equal probability with independent selection(Kenya National Bereau of Statistics, 2022). To generate a sampling frame to choose 25 homes in each cluster, the second stage involved listing houses. However, if a cluster’s size was less than 25, every household in it was sampled. The survey was conducted in over 1691 clusters. The training of data collectors and pretesting of the study instruments were conducted by the Inner-City Fund (ICF), while data was collected between February and July 2022. Interviews were conducted in Swahili or English with all women aged 15 to 49 years who were regular members of the chosen households or who had spent the night in the household before the survey. 11,863 women who were either pregnant or had given birth during the previous five years were included in the study out of the 32,156 women who completed the survey. Authorization to use the dataset was obtained from the MEASURE DHS website (https://www.dhsprogram.com/data/available-datasets.cfm).

Even though the dataset had numerous variables, only those that were applicable to our study were included in the analysis.

### Study variables

#### Dependent/outcomes

The quality of ANC was the study’s main outcome variable. The quality of ANC, a composite variable, was created by combining several binary (yes or no) questions about the mother’s receipt of certain services during ANC. The services included blood and urine sample collection, blood pressure checks, receiving information about danger signs of pregnancy (e.g., bleeding), fetal heartbeat monitoring, breastfeeding counseling, dietary counseling, and the provision of iron supplements (or purchase). If participants obtained all eight ANC services (Yes), it indicated that they had received quality ANC; and vice versa if one skipped one (No) (Atinga & Baku, 2013; Mpoy et al., 2022; Tadesse Berehe & Modibia, 2020).

#### Independent variables

The three categories of covariates considered in the analysis were sociodemographic, obstetric and prenatal-related, and health facility-related, based on the literature and the data available in KDHS (Atinga & Baku, 2013; Mpoy et al., 2022; Tadesse Berehe & Modibia, 2020). Several sociodemographic parameters were considered and incorporated into the analysis. These variables included the following: education levels of the mother and husband (primary, secondary, or tertiary), age in years (15–19, 20–34, 35–49), wealth index in five classes (poorest to richest), place of residence (rural or urban), marital status (single or married), and religion (Christian, Muslim, or others). Region was categorized into Kenya’s eight provinces— Nyanza, Western, Eastern, Coast, Northeastern, Central, Rift Valley, and Nairobi. The size of the household (≤4 or ≥5) was used to measure family composition. Two proxy variables were used to assess maternal autonomy: who heads the household (female or male) and who makes healthcare-seeking decisions for the participant (partner, self, or jointly with another person/partner, or others). The study also considered phone ownership and exposure (yes or no) to mass media, as having access to newspapers, radio, television, and the Internet. The principal component analysis was also used to compute the wealth index from data on household asset ownership (Kenya National Bereau of Statistics, 2022).

In addition, six antenatal and obstetric factors were analyzed, including whether or not the woman is currently pregnant, whether or not she has received information about antenatal care from a community health worker, parity (≤2, 3–4, ≥5), whether or not she has had antenatal visits (≤3, or ≥4), when she had her first ANC visit in months (0–3, 4-6, 7-9), and whether or not the woman was currently pregnant (Kenya National Bereau of Statistics, 2022).

The analysis also included five variables that were associated with the location where ANC was received. The specific place of the ANC (clinics, faith-based organization (FBO), Non-governmental Organizations (NGO), private, or public health facilities) and the people who evaluated the mother during the ANC (midwife, doctor, clinical officer, nurse, and others) were among these variables. As a proxy measure of access to the health facility, the number of minutes spent to access the health facility of birth (≤30, 31-60, ≥61) was included in the study. One additional proxy measure for participant familiarity with the healthcare facility was whether or not they had ever taken contraceptives (Kenya National Bereau of Statistics, 2022).

### Statistical analysis

Data was cleaned and dummy variables were constructed before analysis. For each categorical variable, descriptive statistics such as frequencies were calculated at the univariate level. We employed univariate logistic regression to identify independent variables associated with the quality of ANC. Simple multivariate logistic regression was then used to identify the variables associated with the quality of ANC while controlling for other variables. All variables with P-values less than 0.05 were included in multivariate analysis and 95% confidence intervals were provided for all odd ratios of variables. The data was analyzed using the complex samples package in SPSS (V20), which assisted in taking into consideration the complicated sample design present in DHS data. According to Croft, Marshall, & Allen (2018) and Zou, Lloyd, & Baumbusch (2020), the complex sample package offers reliable parameter estimations since it takes into consideration weighting, sample stratification, and clustering that happened throughout the study participant sampling process. Furthermore, DHS sample weights were imposed on all computed frequencies to mitigate the effects of unequal probability sampling in various strata and guarantee the representativeness of the study outcomes(Croft, Marshall, & Allen, 2018; Zou, Lloyd, & Baumbusch, 2020). Using a variance inflation factor (VIF) of less than 10 as a cutoff, multi-collinearity was also evaluated among all the predictor variables in the model (Croft et al., 2018; Zou et al., 2020). All the predictors fell below the threshold.

### Ethical consideration

The Institutional Review Board of the Inner-City Fund (ICF) granted ethical approval for the 2022 KDHS. Whereas the Kenya National Bureau of Statistics carried out the study in collaboration with other development partners. Since this study is based on secondary data from the KDHS that is publicly available, no ethical approval was required for its analysis, however, MEASURE DHS provided authorization to use the KDHS datasets (https://www.dhsprogram.com/data/available-datasets.cfm). Both from human participants and from legally appointed representatives of minor participants, written informed consent was acquired.

## Results

### Demographic characteristics of the study participants

In total, 11,863 women who were pregnant or had given birth within the five years before the survey were included in this analysis (Table 1). The majority identified themselves as being from the Central, Eastern, Rift Valley, Nyanza, and Nairobi provinces (76.7%), were between the ages of 20 and 34 (74.4%), and 61.4 % lived in rural areas. The majority of participants were married (80.2%), had only completed their primary (or none) education (44.8%), and identified as Christians (88.5%). Fifty-seven percent (57.3%) were employed, and nearly forty-four percent (43.9%) belonged to the richest quintiles. Furthermore, 91% of the participants’ husbands were employed and the majority had at least a secondary education (55.9%). The majority made decisions to seek health care services jointly with their partner or another person (44.9%), and they lived in male-headed households (71.5%), with their partners (83.2%), and with more than five household members (79%). A significant proportion of the participants were exposed to mainstream media, which included newspapers (17.2%), television (38.4%), the internet (47.9%), and radio (74.3%). Furthermore, 81.1 % of the participants were mobile phone owners. Although 94% were not pregnant, most participants (91.2%) wanted/desired to have their most recent pregnancy and had given birth to utmost two children or had utmost two children currently alive (53.9%).

**Table 1:**
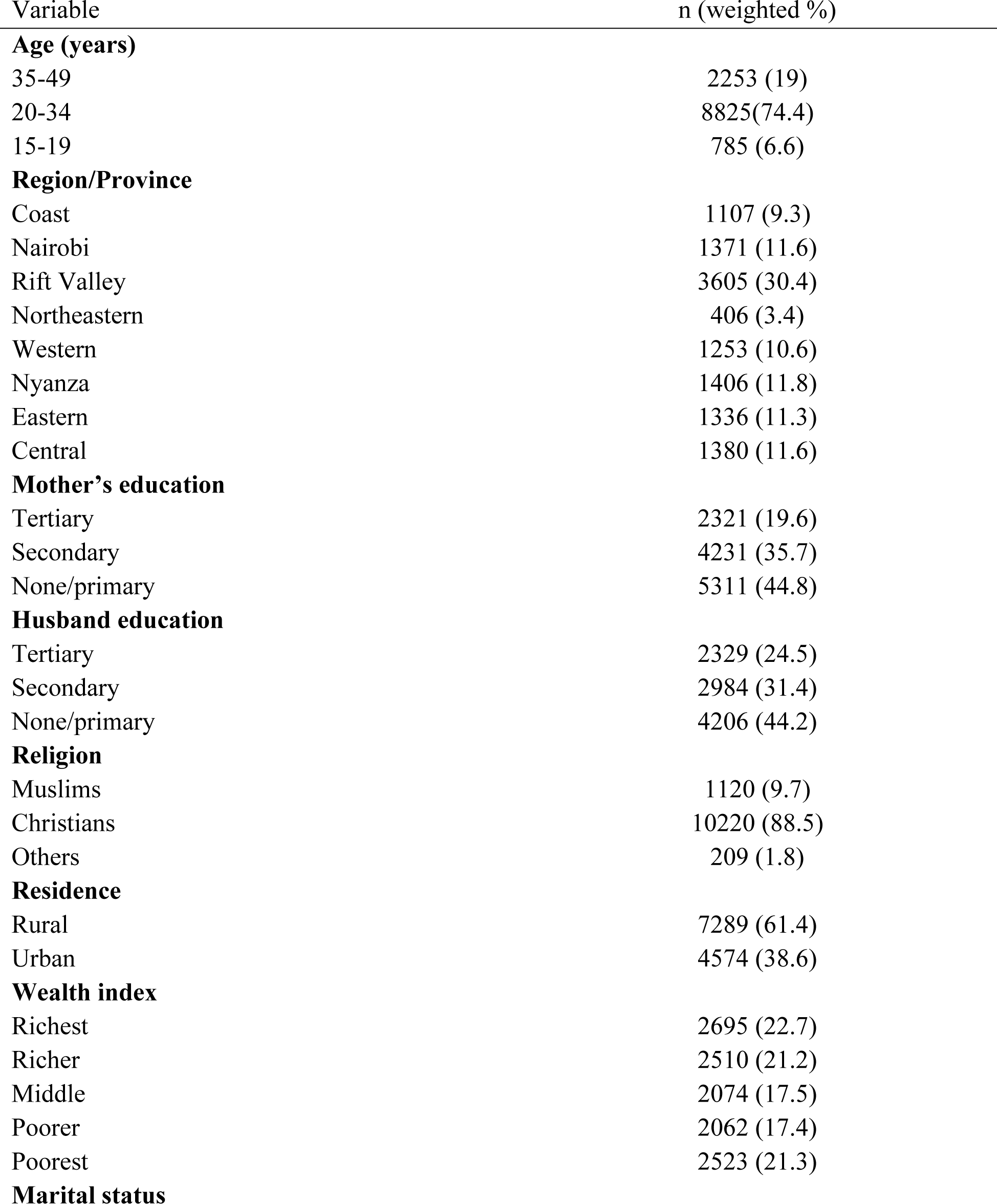
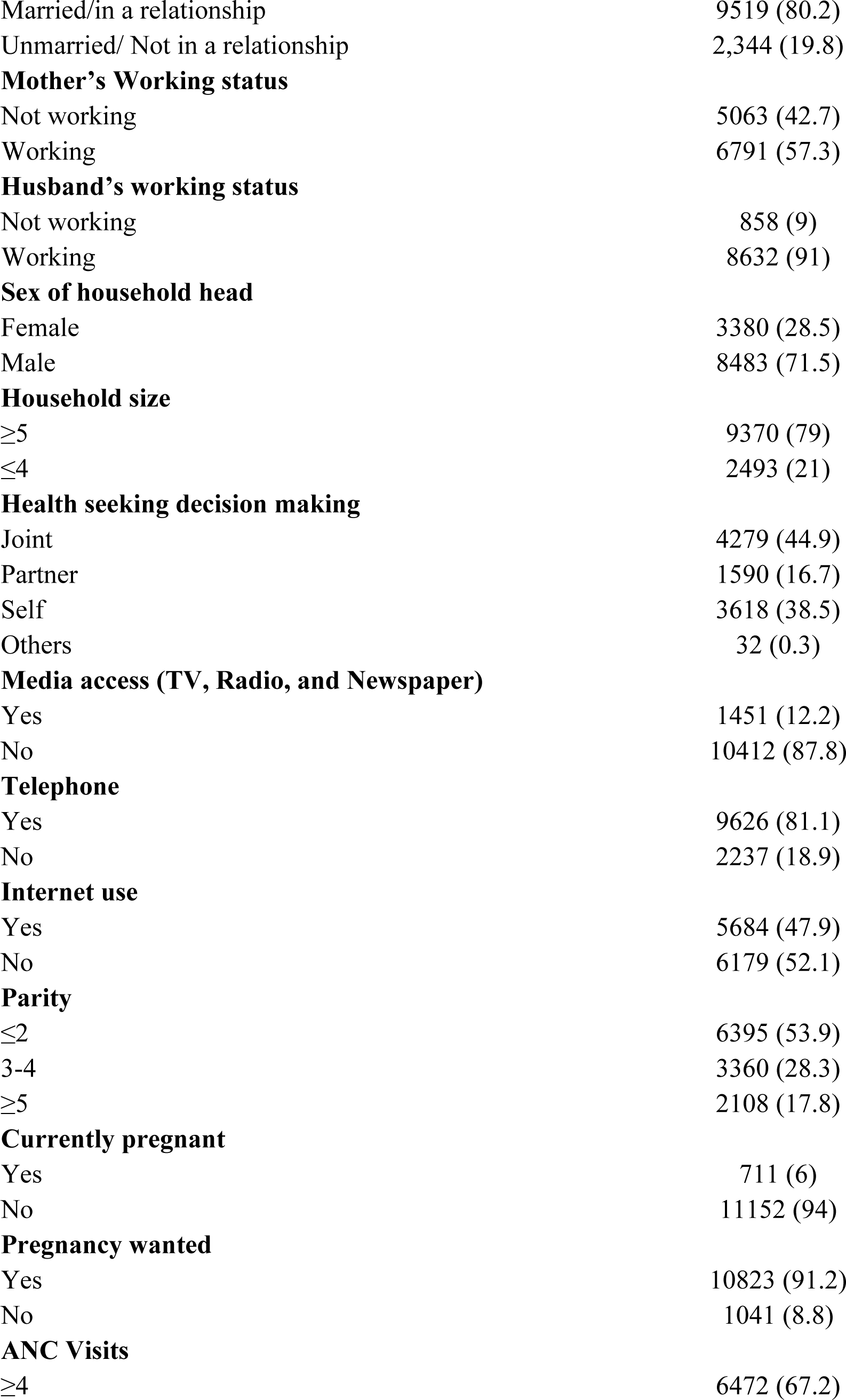
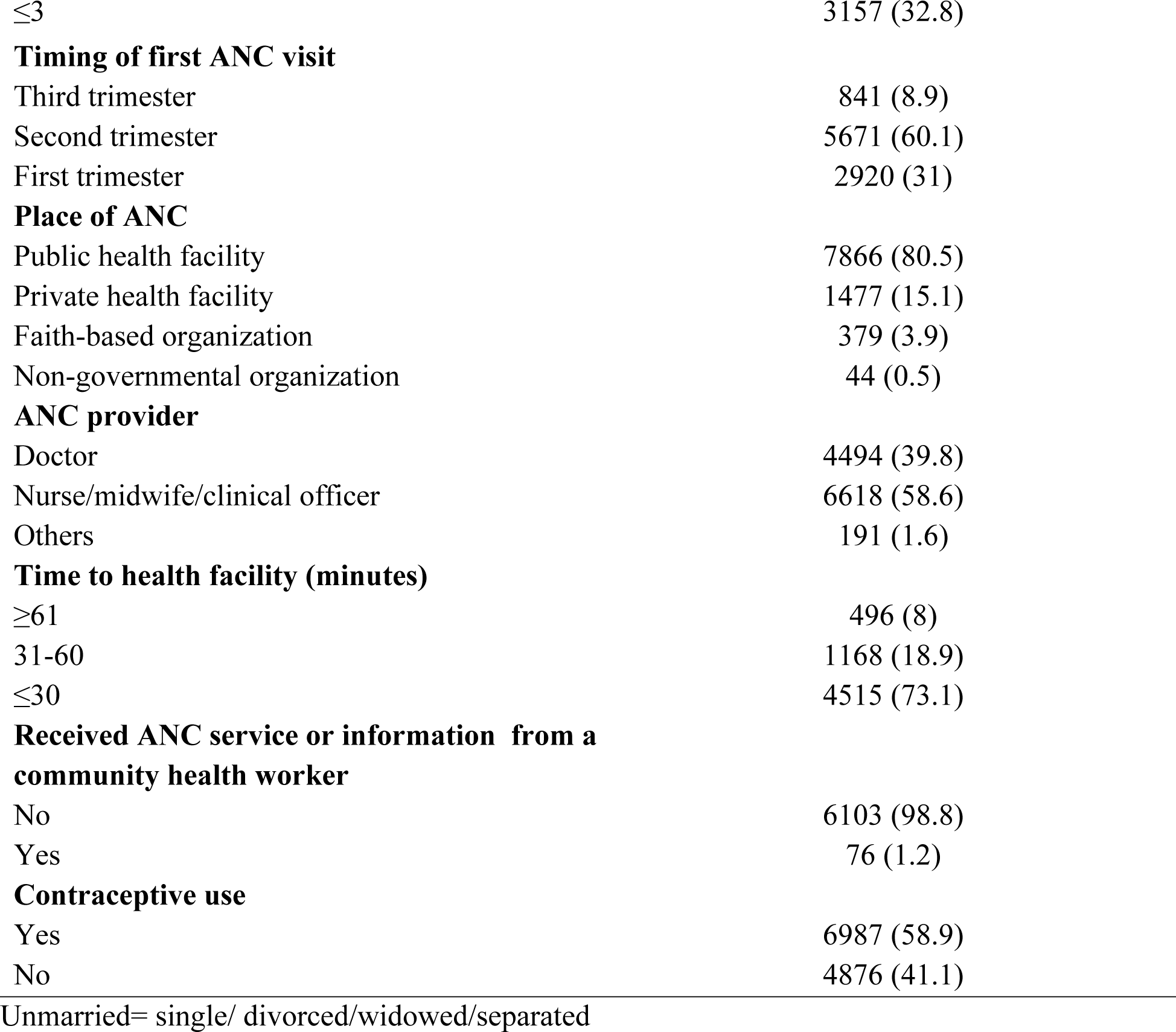
Demographic characteristics of the study participants

The majority (60.1%) had attended their first prenatal appointment in the second trimester, and 67.2% had attended at least four antennal appointments overall from public (83.4%) and private (15.7%) healthcare facilities. Most had received ANC from doctors (46.7%) and midwives/nurses/clinical officers (68.7%). Merely 1.2% of the mothers reported learning about ANC from a community health professional. Furthermore, most mothers had used contraceptives (58.9%) and had walked (67.7%) to health facilities for ANC, for a maximum travel time of 30 minutes (73.1%).

### Quality of Antenatal Care

Overall, 61.2% of the study participants received quality antenatal care (**Table 2**). Among the most used ANC services, 98.2% of the participants had their blood pressure taken, followed by 97.7 % of the participants whose fetal heartbeat was monitored, and 97.1% and 95.9% had their blood and urine samples taken off, respectively. Meanwhile, 92.2% of the participants also received iron tablets. On the other hand, fewer participants received nutritional counseling (84.4%), breastfeeding counseling (82.4%) and counseling about the danger signs of pregnancy (77.0%).

**Table 2:** Component of antenatal care received by study participants.

| Variable | n (weighted %), 95%CI |
| --- | --- |
| **The overall quality of Antenatal care** | **61.2 (59.7-62.6)** |
| Blood pressure taken | 98.2 (97.9-98.4) |
| Urine samples taken | 95.9 (95.4-96.4) |
| Blood samples taken | 97.1 (96.7-97.5) |
| Fetal heartbeat monitored | 97.7 (97.4-98.1) |
| Nutritional counselling | 84.4 (83.5-85.4) |
| Breastfeeding counselling | 82.4 (81.3-83.4) |
| Advise on danger signs of pregnancy (bleeding) | 77.0 (75.7-78.3) |
| Given/bought iron tablets | 92.2 (91.4-92.8) |

### Factors associated with the quality of antenatal care

The factors associated with the quality of antenatal care in univariate and multivariate logistic regression analysis are summarized in **Table 3**. Older mothers (aged 20–34) had a 1.82 (95%CI: 1.15-2.87) times higher likelihood of receiving quality ANC when compared with younger mothers (15–19 years old). Participating mothers who had attended 4 or more ANC visits were 1.42 (95%CI: 1.14-1.79) times more likely to receive quality ANC than those who attended 3 or less visits. Comparing participants with and without media access, those with media access were 1.47 (95%CI: 1.06-2.03) times more likely to receive quality ANC. Furthermore, the likelihood of receiving quality ANC was 1.93 (95%CI: 1.21-3.08) and 1.44 (95%CI: 1.01-2.06) times higher for participants in the richest and richer quintiles, respectively, than for those in the poorest quintile.

**Table 3:**
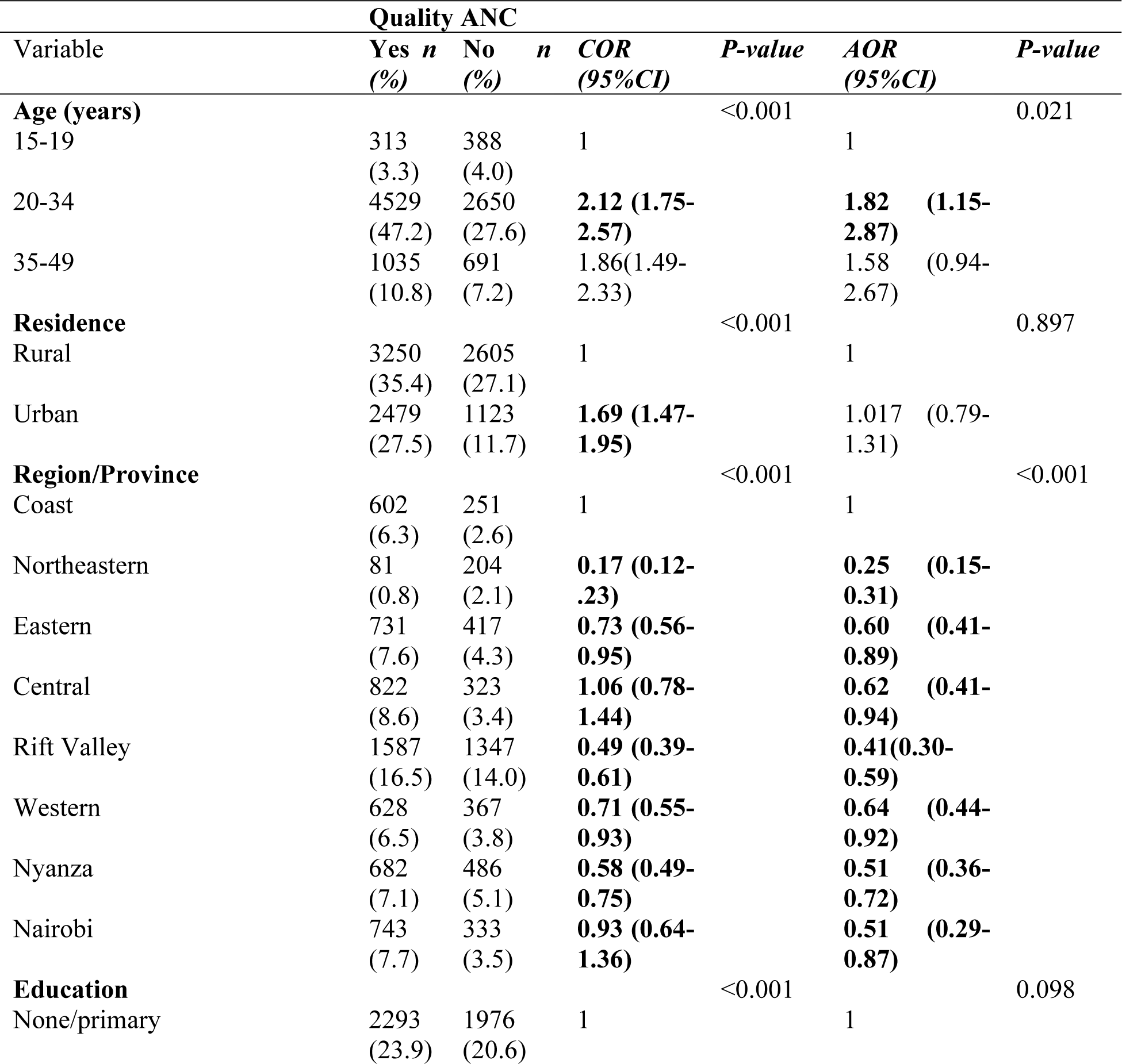
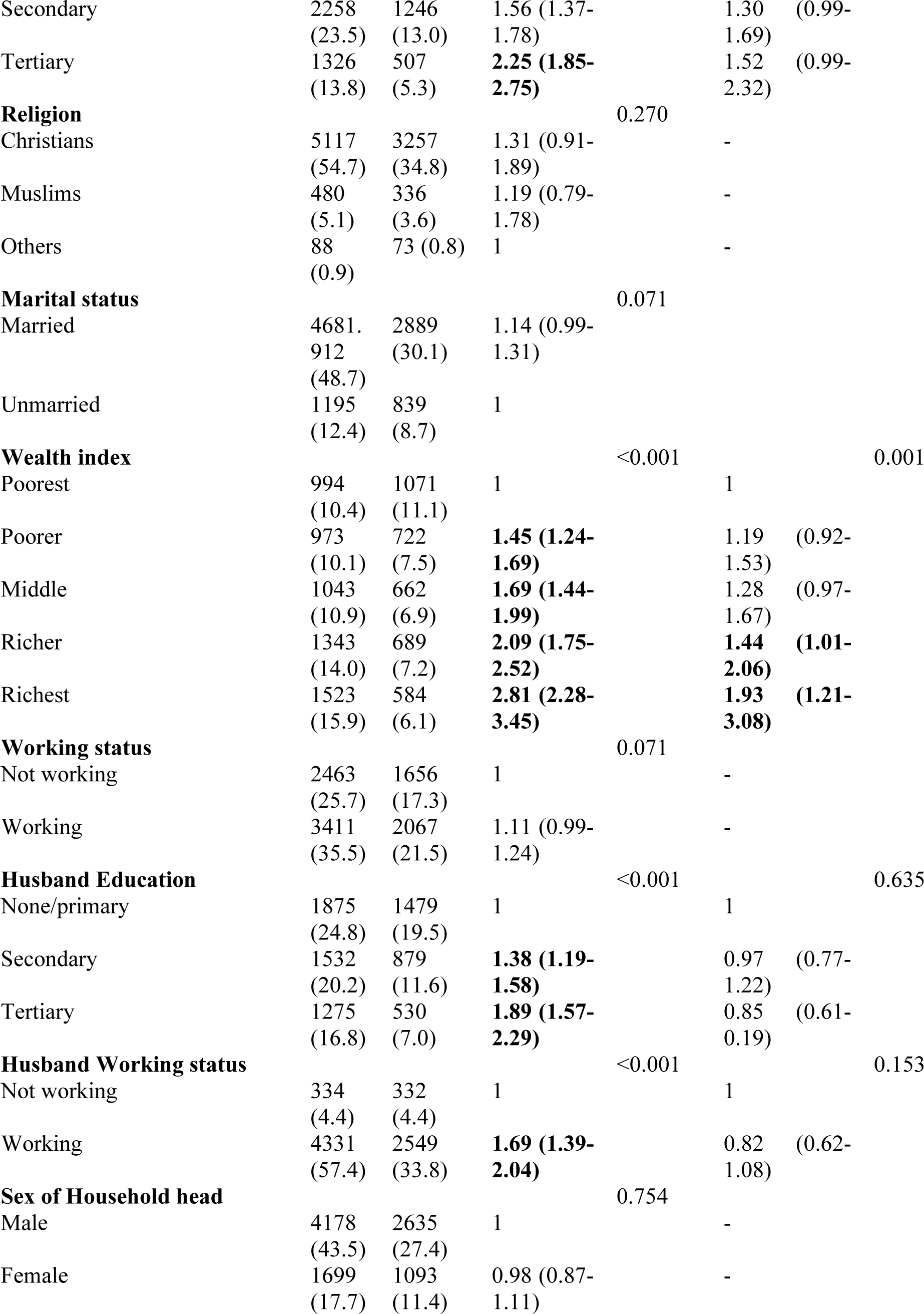
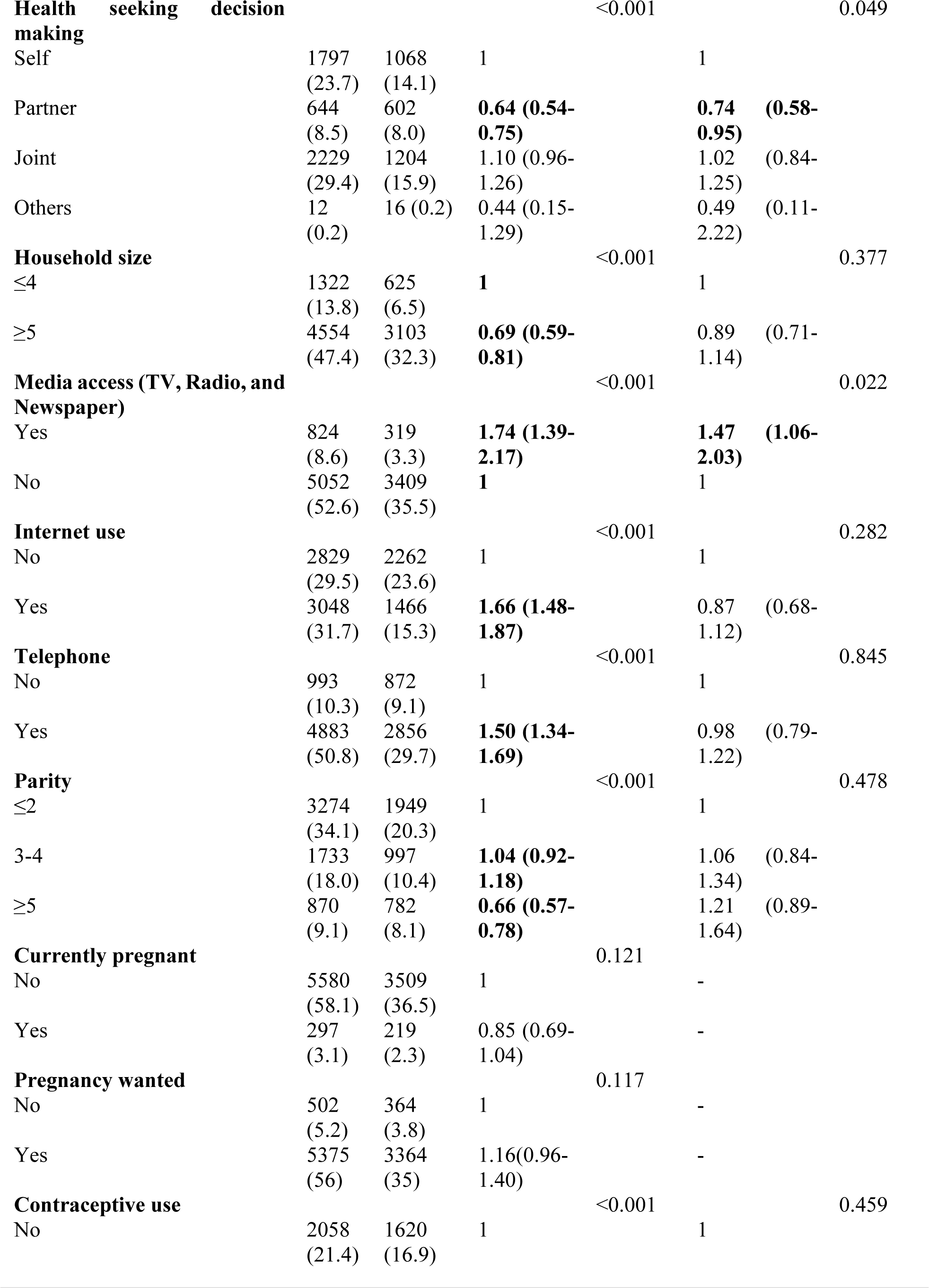
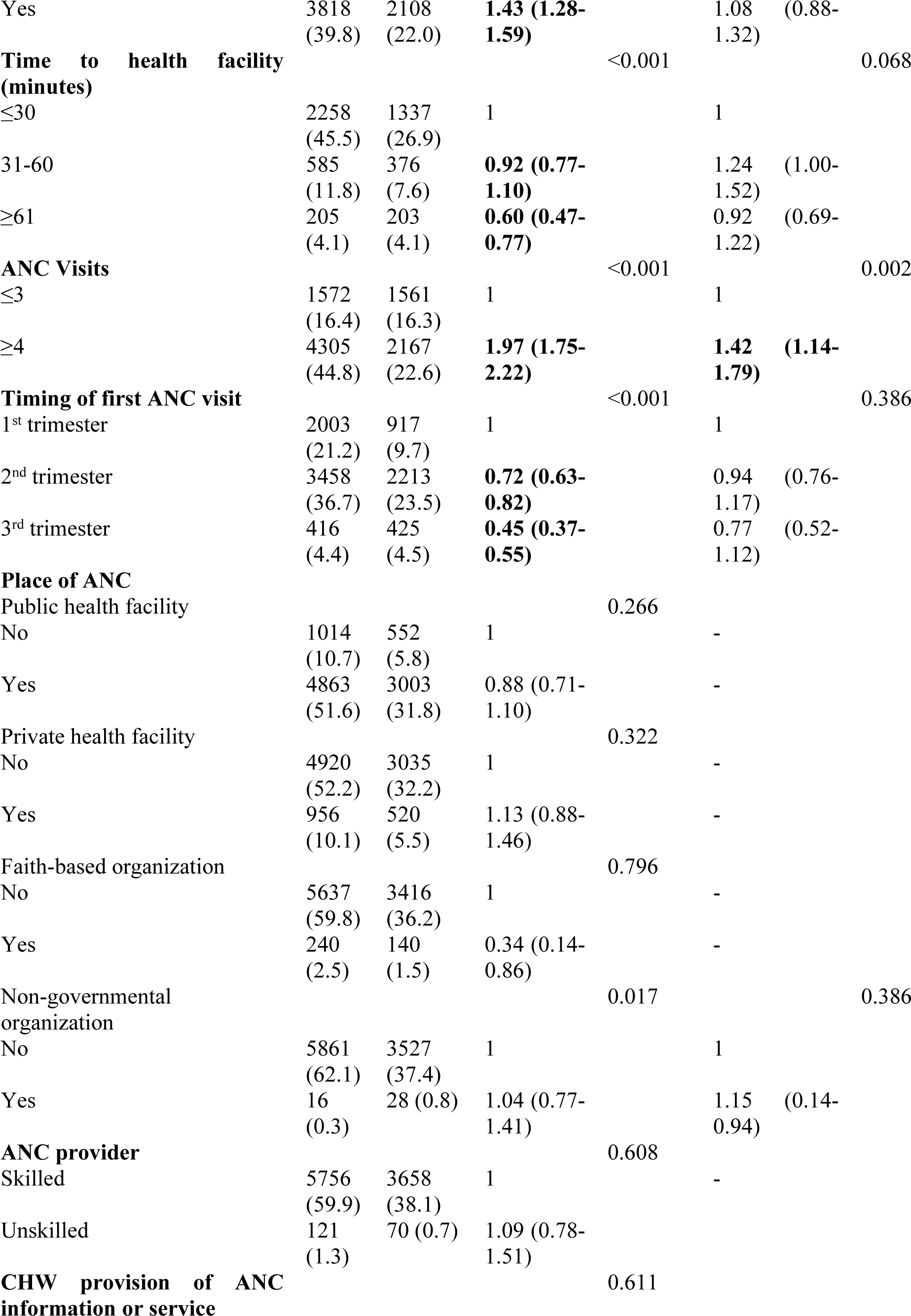
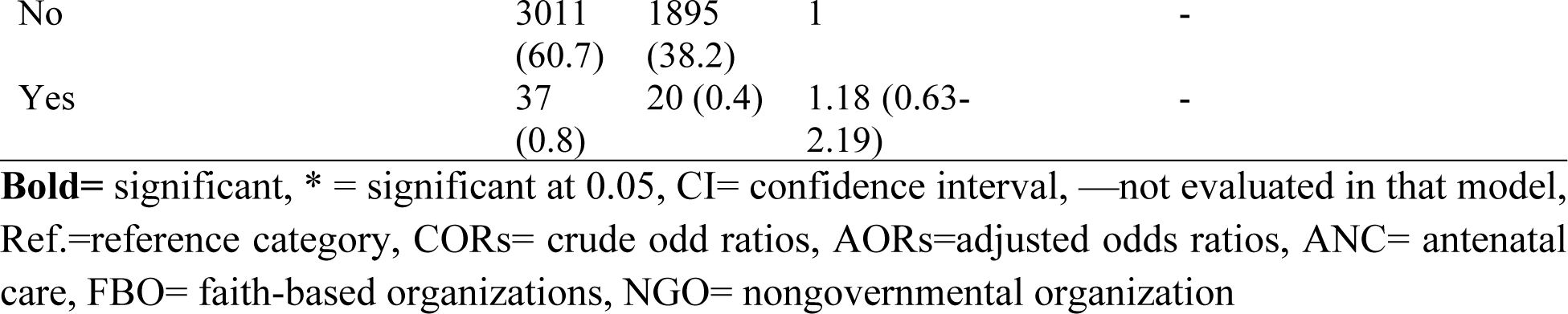
Factors associated with the quality of antenatal care among women aged 15-49 years.

On the contrary, the odds of receiving quality ANC were 0.25 (95%CI: 0.15-0.31) to 0.64 (95%CI: 0.44-0.92) times lower for participating mothers from all other Kenyan regions than for those from the coastal region. Participants whose husbands or partners made decisions for them to seek healthcare, compared with those who made decisions independently were 0.74 (95%CI: 0.58-0.95) times less likely to receive quality antenatal care.

## Discussion

This study assessed the factors associated with the quality of ANC in Kenya using the 2022 demographic health survey. The overall prevalence of quality ANC among study participants was found to be 61.2%. This is higher than the prevalence of the quality of ANC from Ethiopia at 31.38% (Tadesse Berehe & Modibia, 2020), Rwanda at 13.1% (Sserwanja et al., 2022) and Nigeria at 45% (Idowu, Israel, & Akande, 2022). The difference between our study and the literature may be attributed to the differences in health policies, health facility standards, and ANC implementation challenges in various countries (Idowu et al., 2022) (Ajibola Idowu, 2022 & MoH Kenya 2021). Additionally, differential efforts and resources put into maternal health services, as well as differences in socio-demographics across countries may explain the differences in findings between this study and the literature (Zhao et al., 2023, Kare, 2021; Tuncalp et al., 2016).

In the multivariate analysis, age was significantly associated with the quality of ANC. Women aged 20-34 years were more likely to receive quality ANC than younger women aged 15-19 years. This finding is consistent with studies conducted in Bangladesh and sub-Saharan Africa, which reported that younger women were less likely to receive quality ANC (Akter, 2023 & Garcia et al., 2021). Additionally, women aged 20-34 years are more likely to have better knowledge and understanding of the benefits of quality ANC due to increased awareness and proactive engagement with the healthcare system, increasing their demand and use for such services (Smith et al., 2020).

In terms of region, there were significant variations in the quality of ANC across different provinces in Kenya. For example, the Rift Valley region had lower odds of receiving quality ANC, while the Northeastern region had the lowest odds compared to the Coastal region. The coastal region is in proximity to urban centers and tourist areas of Kenya with better healthcare facilities (Phiri et al. 2014). The findings may indicate the presence of regional differences and variations in the provision of ANC, with some regions facing challenges such as shortages of skilled healthcare providers, inadequate infrastructure, and limited access to essential resources (Mwangome et al., 2020). These regional disparities highlight the importance of addressing geographical inequities in healthcare delivery.

The Wealth Index was significantly associated with quality ANC utilization. The richest participants had higher odds of receiving quality ANC compared to the poorest participants. Similar studies in Nigeria reported that the wealthiest respondents were more likely to adequately use ANC (Fagbamigbe et al., 2017& Feng et al., 2021). Richest women do not have the challenges of accessing the money necessary for transport to health facilities and accessing more sophisticated healthcare (Fagbamigbe et al., 2017). Empowering Kenyan women through adequate employment/income generating activities should be paramount in policies targeted at optimizing quality ANC utilization.

Health-seeking decision-making was also associated with quality ANC utilization. Participants whose partners made decisions for them to seek healthcare were less likely to receive quality ANC compared with those who had joint decision-making. This finding may be partly explained by a qualitative study conducted in Malawi which found that most men termed pregnancy and ANC as a women’s issue, thus perceiving decision-making around ANC as a low priority (Kennedy Machira1, 2023). On the other hand, women’s autonomy in decision-making has a positive effect on ANC service use (Awoleye et al., 2018 &Haider et al., 2017). Therefore, it is recommended to implement interventions that promote shared decision-making and autonomy in healthcare-seeking behaviors among pregnant women and their partners.

Access to media was significantly associated with quality ANC utilization. The findings are similar to studies in Uganda (Bbaale, 2011) and Bangladesh (Akter et al., 2023) that established a significant correlation between media exposure and the quality of ANC. This was because media exposure gave mothers visual and audio access to health-related information, which eventually improved access to healthcare and demand for ANC services (Ndugga, Namiyonga, & Sebuwufu, 2020; Zhao, Zhang, Xiao, He, & Tang, 2023). We recommend that more reviews are conducted to establish the most effective media for quality ANC utilization.

In this study, the number of ANC visits attended was significantly associated with the quality of ANC utilization. Women who had attended 4 or more ANC visits were more likely to receive quality ANC compared with those who had attended less or equal to 3 visits. This finding is similar to an Ethiopian study by Kare et al. (2021) that found that pregnant women who had visited hospitals for ANC four or more times had higher odds of having quality ANC. Attending ANC more than 4 times increases the chances of obtaining multiple ANC services including identifying complications and risky behaviors during pregnancy and is a very important indicator of quality ANC as stated by WHO (2016).

### Limitations and strengths

We utilized the most recent data from the 2022 Kenya Demographic and Health Survey, with a large sample size and rigorous data collection protocols making the results of our study generalizable to Kenya and sub-Saharan Africa. However, the study is based on retrospective data provided by the survey respondents, which may be subject to recall bias. In addition, the cross-sectional design of this study infers association and not causality.

## Conclusion

The study revealed that 61.2% of the mothers had received quality ANC. The study also reported several factors associated with quality ANC, age, region, wealth index, health-seeking decision making, access to media (TV), and ANC visits. It is recommended to implement interventions that promote shared decision-making and autonomy in healthcare-seeking behaviors among pregnant women and their partners. The regional disparities highlight the importance of addressing geographical inequities in healthcare delivery. We recommend that more reviews are conducted to establish the most effective media for quality ANC utilization.

## Acknowledgments

We are grateful that the data used in this investigation were made available by the Demographic Health Survey program.

## Availability of data and materials

The data set used for this study is openly available upon obtaining permission from the MEASURE DHS website (URL: https://www.dhsprogram.com/data/available-datasets.cfm). However, authors are not authorized to share this data set with the public however anyone interested in the data set can seek it with written permission from the MEASURE DHS website (URL: https://www.dhsprogram.com/data/available-datasets.cfm).

## Authors’ contributions

**Concept and proposal development:** John Baptist Asiimmwe, Lilian Nuwabaine,

**Data analysis:** John Baptist Asiimmwe, Lilian Nuwabaine, Joseph Kawuki, Quraish Sserwanja, Angella Namulema, Mathius Amperiize, Earnest Amwiine

**Writing the original draft:** John Baptist Asiimmwe, Lilian Nuwabaine, Joseph Kawuki, Quraish Sserwanja, Angella Namulema, Mathius Amperiize, Earnest Amwiine

**Writing-review, and editing:** John Baptist Asiimmwe, Lilian Nuwabaine, Joseph Kawuki, Quraish Sserwanja, Angella Namulema, Mathius Amperiize, Earnest Amwiine

## Ethics approval and consent to participate

The study protocol is carried out following pertinent criteria, and high international ethical standards are guaranteed during the MEASURE DHS surveys. The ICF Institutional Review Board assessed and approved the protocol for the 2022 KDHS survey. Both from human participants and from legally appointed representatives of minor participants, written informed consent was acquired.

## Consent for publication

This is not applicable in this study.

## Abbreviations

AOR: Adjusted Odds Ratio
ANC: Antenatal Care
EA: Enumeration area
CI: Confidence Interval
COR: Crude Odds Ratio
DHS: Demographic Health Survey
KDHS: Kenya Demographic Health Survey
OR: Odds Ratio
SPSS: Statistical Package for Social Science
VIF: Variance Inflation Factor
WHO: World Health Organization

